# An approach to quantifying the interaction between behavioural and transmission clusters: the case of Hepatitis C virus infections in HIV positive men who have sex with men

**DOI:** 10.1101/2021.08.22.21261697

**Authors:** Luisa Salazar-Vizcaya, Katharina Kusejko, Huldrych F Günthard, Jürg Böni, Karin Metzner, Dominique Braun, Dunja Nicca, Enos Bernasconi, Alexandra Calmy, Katharine EA Darling, Gilles Wandeler, Roger D Kouyos, Andri Rauch, the Swiss HIV Cohort Study

## Abstract

We hypothesize that patterns of sexual behaviour play a role in the conformation of transmission networks. If that was the case, behavioural grouping might in turn correlate with transmission networks and have the potential to predict proximity in viral phylogenies. To address this hypothesis, we present an intuitive approach for quantifying interactions between clusters of sexual behaviour along a virus phylogeny. Data from the Swiss HIV Cohort Study on condom use and incident Hepatitis C virus (HCV) sequences served as proof-of-concept. A strict inclusion criteria contrasting with relatively low HCV prevalence hindered our ability to identify significant relationships. This manuscript intends to serve as guide for studies aimed at characterizing interactions between behavioural patterns and transmission networks. Large transmission networks such as those of HIV or COVID-19 are prime candidates for applying this methodological approach.

## INTRODUCTION

Sexual behaviour has changed among men who have sex with men (MSM) over the last two decades. In particular, condom use might have been influenced by changes in recommendations related to emerging scientific knowledge, availability of social media and the widespread use of stimulating substances for sex [1, 2]. Such behavioural changes demonstrated heterogeneous trends [3-6]. Groups defined by similar patterns of change in sexual behaviour could reflect similar adaptation mechanisms to changes in information capable of influencing risk perception. For instance, the popularization of the U = U message (undetectable equals untransmissible) [7, 8] [9] preceded by the Swiss Statement [10] might have only reduced condom use among some persons living with HIV during sexual intercourse with partners of HIV negative or unknown status. However, the probability that such changes in risk behaviour occur, may vary across individuals and over time. For example, we recently identified clusters of sexual behaviour with distinct time trends in condom use during anal intercourse [6]. We hypothesized that these behavioural clusters may play a role in the conformation of viral transmission networks and vice versa. If this was the case, behavioural groups inferred from temporal changes in sexual practices might in turn help predict proximity in transmission networks.

Clear characterizations of the relationships between sexual behaviour and transmission networks may help identify targets for efficient treatment and behavioural interventions. We propose an approach to help quantify such relationships. Associations between condom use and incident infections with hepatitis C virus (HCV) have been shown previously [11]. Here we investigated HCV infections among HIV-positive MSM for proof of concept using behavioural and HCV sequence data from the Swiss HIV Cohort Study (SHCS) and focusing on analysing intersections between clusters of sexual behaviour and clusters of HCV transmission. We used these intersections to infer interactions between clusters of sexual behaviour.

## RESULTS

This analysis studies the overlap between two types of clusters previously inferred: 1) Behavioural Clusters (***BCs***), and 2) Transmission clusters (***TCs***). The methods section includes a graphical illustration of this process.

1. ***BCs:*** The characteristics of the considered behavioural clusters have been described in detail before [6] [12]. In brief, a pattern recognition method led to clustering of MSM according to their history of condomless anal intercourse with non-steady partners (nsCAI) as recorded by the SHCS. We termed the resulting groups, behavioural clusters (*BC*). The top four BC of this classification revealed evidently divergent trajectories of nsCAI [6] [12]. Here we also studied the two top *BC*. In order to cover the broader behavioural spectrum, the present study included an additional group, comprising MSM who have never reported nsCAI (*BC*_*0*_). We referred to the resulting sets of behavioural clusters as the *full classification* (top 4 + *BC*_*0*_) and the *three clusters classification* (top 2 + *BC*_*0*_). Size, nsCAI trajectories over time for both classifications are briefly summarized in **Figures 1A** and **1B**. **Figure S1** shows the nsCAI trajectories over time for the two no nil clusters in the *three clusters classification*. The analogous trajectories for the *full classification* were reported in [6].
2. ***TCs:*** These clusters have been described previously [13]. They are 11 and based based on a phylogenetic tree including Swiss and foreign HCV genetic sequences comprising a segment of the NS5B region of the virus. *TCs* were defined as monophyletic trees with bootstrap support value larger than 70%.

**Figure 1.**
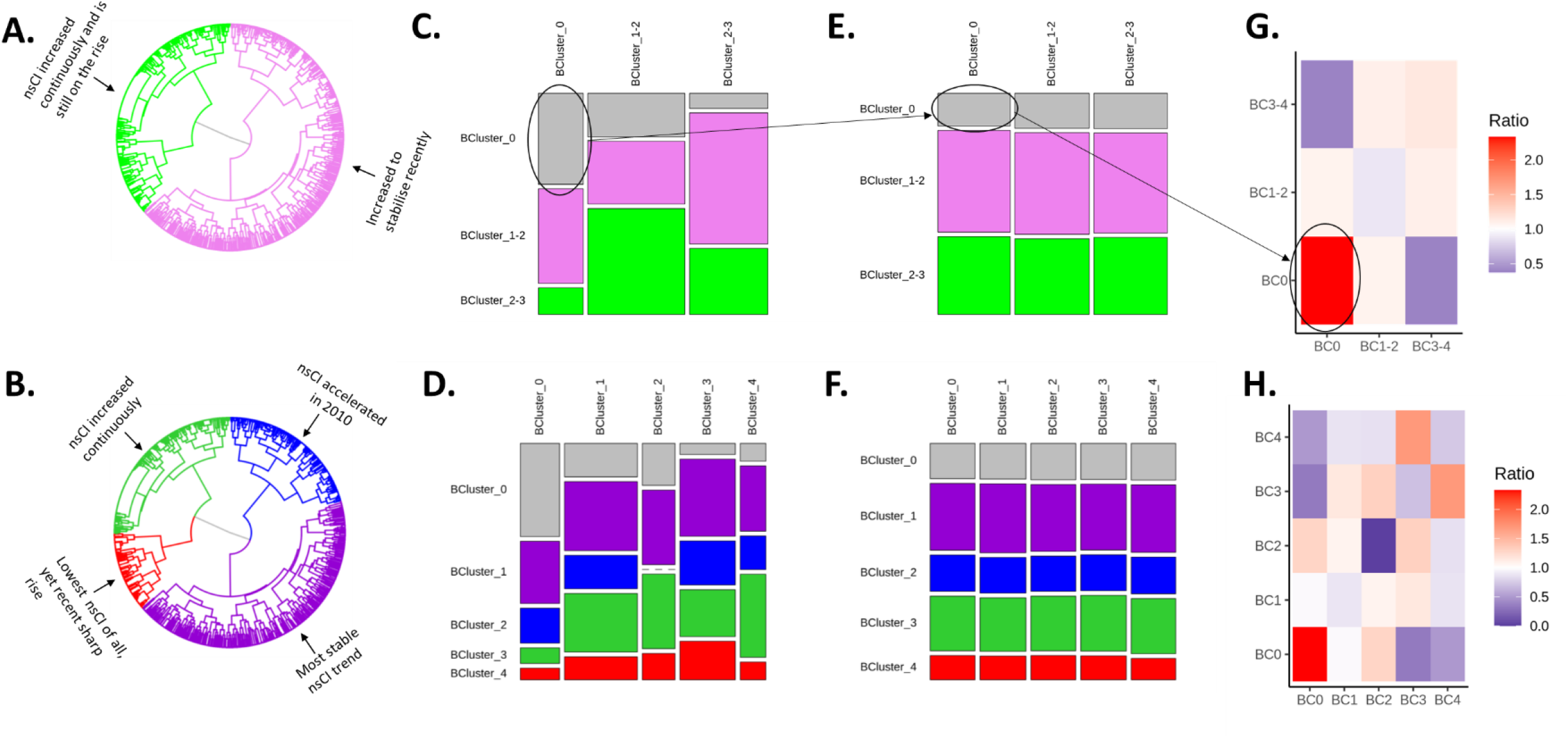
Interactions between behavioural clusters mediated by HCV transmission clusters. **Behavioural clusters diagrams** for: **A**. The Three clusters classification, and **B**. The Full classification. The cluster without nsCAI (BC0) is not shown since it does not emerge as an outcome of the clustering algorithm. **Mosaic plots for observed** interactions between behavioural clusters mediated by HCV transmission clusters for: **C**. the three clusters classification, and **D**. the full classification. (Hint: vertical length of rectangles reflects the relative magnitude of the estimate) **Mosaic plots for expected interactions** between behavioural clusters mediated by HCV transmission clusters for: **E**. the three clusters classification, and F. the full classification. **Ratios between observed and expected mixing amongst members of behavioural clusters** for: **G**. The three clusters classification and the **H**. The full classification **Hint:** The grey rectangle (representing the interaction between members of BC0) in the observed diagram (**C**) is taller than that in the expected diagram (E). This indicates that members of BC0 were more likely than expected to meet other members of the BC0 in a transmission cluster. The corresponding cell in G is the most intense since it is also the strongest overrepresentation we found.

### Intersections: Mapping behaviour on transmission

We mapped membership to one of five *BC*s into each *TC*. Out of 4222 MSM with identified behavioural cluster membership, and 66 in the HCV phylogeny, 36 mapped to one of the *TCs i*.*e*., had a HCV sequence located in a transmission cluster. Six members of cluster *BC*_*0*_, 11 of *BC*_*1*_, 6 of *BC*_*2*_, 9 of *BC*_*3*_ and 4 of *BC*_*4*_ mapped to any of the *TCs*.

### Are MSM who shared a behavioural cluster likely to also share transmission cluster?

#### Full classification

The odds ratio for sharing both behavioural and transmission clusters versus sharing behavioural but not transmission clusters after 10,000 iterations was 0.97 (p-value 0.61).

#### Sample size and power considerations

This analysis included 107 independent pairs of men sharing transmission cluster and 433 not sharing transmission cluster. To achieve a power of 70% at a level of significance of 0.05 for this ratio would require more than 98000 men in each of these two groups.

### Potential influence of behavioural patterns on HCV transmission clustering

**Figures 1C-F** depict, for visual comparison, the observed (**C** and **D**) and expected (E and **F**) interactions between behavioural clusters mediated by HCV transmission clusters, for the *full* and the *three clusters* classifications, respectively. For instance, **figures 1C and 1E** indicate that the estimated interaction between members of *BC*_*0*_ was stronger than expected, suggesting that members of *BC*_*0*_ are prone to share transmission cluster. By contrast, the estimated interaction between members of clusters *BC*_*0*_ and *BC*_*3*_ was weaker than expected. Expected interactions constitute our null hypothesis and were assumed to only depend on the overall frequency of behavioural clusters along the HCV phylogeny *i*.*e*., random grouping, so that interaction with larger clusters is more likely.

**Figure 1G** depicts the ratios between observed and expected interactions amongst members of the behavioural clusters along the HCV phylogeny for the *three clusters classification*. For instance, the largest resulting overrepresentation was found among members of *BC*_*0*_ (2.3, p-value: 0.45), while the most prominent no nil underrepresentation was found to occur between members of *BC*_*0*_ and members of *BC*_*3-4*_ (0.37, p-value: 0.54). Analogously, **Figure 1H depicts** the respective ratios for the *full classification*. This ratio was also maximal for interactions among members of *BC*_*0*_, while we recorded no interactions among members of *BC*_*2*_. **Sample size and power considerations:** We estimated that overrepresentations and underrepresentations comparable to the strongest found in this proof-of concept could require 3 and 4 times as many mapped men (108 and 144 versus 36 available) respectively to achieve a power of 70% at a level of significance of 0.05.

## DISCUSSION

We present an intuitive approach for quantifying associations between independently obtained behavioural and transmission clusters, which could help understand whether and how behavioural patterns relate to transmission networks. We used behavioural clusters based on condom use with non-steady partners and incident hepatitis C virus infections for proof-of-concept. Lack of power hindered our possibilities of obtaining conclusive outcomes for this proof of concept. This manuscript intends to serve as guide for further, powered studies aimed at characterizing such associations.

This is the first study to quantify associations between clusters of sexual behaviour and clusters of transmission derived from a viral phylogenie. We did this based on a simple, reproducible framework suitable for analogous and more general studies. Phylogenetic information bigger than that available for our proof-of-concept is however necessary to reach conclusive results (e.g. on a more prevalent, densely sampled infection, such as HIV).

Our proof-of-concept relied on HCV infections, which resulted in a low-density mapping on the behavioural clusters, a risk that arises from analysing a setting with small population: while the HCV subtype 1a phylogenetic tree we used, reached a sampling proportion above 60% (note that 60 sequences potentially relevant for our proof of concept were located in well defined transmission clusters) [13], only 36 patients meet the criteria to be included in this comparison.

Grounding an analysis of this nature on HCV among MSM in a geographical region with larger, denser transmission networks could reach the needed power and carry key advantages. Firstly, increased HCV transmission and changes in sexual behaviour occurred simultaneously during the last years. Secondly, behavioural clusters depict behaviour after HIV diagnosis, and HCV infections in MSM have been most often observed to occur after HIV infection. Thirdly, the incidence of HCV has been on the rise among MSM in industrialized countries since the beginning of the 2000’s [14-16], with recent incipient signs of decline presumably attributable to the scale-up of direct-acting antivirals [17, 18].

That viral proximity and similar sexual behaviour could be related has been hinted by recent studies on drivers of HIV transmission among MSM in Switzerland [19, 20]. Kusejko and co-authors found that condomless sex was more common among HIV phylogenetic neighbours of MSM who did not use condoms [20]. Such type of relationships may be mediated and at least partially explained by social networking through the exchange of information. The authors of a literature review of MSM networks research studies concluded that norms, attitudes, and levels of exposure to HIV transmission were similar among members of the same social network [21]. Other studies involving behaviour and transmission focused on behaviour as predictor of seroconversion. For instance, a study in Amsterdam identified three typical trajectories of anal intercourse with occasional partners among MSM over their life’s course and assessed the risk of HIV seroconversion across trajectories [22].

In summary, this manuscript presents a methodological approach to assess whether records on sexual behaviour could aid inference on transmission networking. For instance, it might be possible to detect (or dismiss) assortativity (preferential interaction with members of the same group) along transmission networks. Assortativity could imply that MSM with similar behavioural patterns would also likely share transmission networks. The prevalence of HCV infection is relatively low [23] and international transmission prominent [13]. We were therefore limited to small numbers, underscoring the need for larger transmission networks, as they would enable dense mapping. Large transmission networks such as those of HIV or COVID-19 are prime candidates for applying this methodological approach.

## METHODS

### The Swiss HIV cohort study

The SHCS (www.shcs.ch) is a nationwide prospective cohort that routinely collects behavioural, laboratory, and clinical data from HIV positive persons aged ≥ 16 years since 1988. The cohort records individual data at study entry and in bi-annual follow-up visits. We estimate that more than 80% of all MSM currently diagnosed with HIV in Switzerland are followed in the cohort [24, 25].

The SHCS has been approved by the ethics committee of the participating institutions (Kantonale Ethikkommission Bern, Ethikkommission des Kantons St. Gallen, Comite departemental d’ethique des specialites medicales et de medicine communataire et de premier recours, Hôpitaux Cantonale de Genève, Kantonale Ethikkommission Zürich, Repubblica e Cantone Ticino—Comitato Ethico Cantonale, Commission cantonale d’étique de la recherche sur l’être humain, Canton de Vaud, Lausanne, Ethikkommission beider Basel for the SHCS and Kantonale Ethikkommission Zürich for the ZPHI). Written informed consent had been obtained from all participants. All methods were carried out in accordance with Swiss federal guidelines and regulations.

This analysis involved two types of clusters previously inferred: 1) Behavioural Clusters (***BCs***); and 2) Transmission clusters (***TCs***).

1. ***BCs:*** The characteristics of the considered behavioural clusters have been described in detail before [6] [12]. In brief, a pattern recognition method led to clustering of MSM according to their history of condomless anal intercourse with non-steady partners (nsCAI) as recorded by the SHCS. NsCAI trends in the top four clusters of this classification revealed divergent trajectories of nsCAI. We refer to these as behavioural clusters (*BCs*).
2. ***TCs:*** HCV transmission clusters have also been described previously [13]. They are based on a phylogenetic approach including Swiss and foreign HCV genetic sequences comprising the NS5B region of the virus. *TCs* were defined as monophyletic subtrees with bootstrap support value ≥ 70%.

### Intersections: Mapping behaviour on transmission

We mapped membership to one of the *BCs* into one of the *TCs*. In accordance with the inclusion criteria for the primary two studies [6, 13], we considered MSM enrolled in the Swiss HIV Cohort Study (www.shcs.ch) with HCV sequences located in HCV transmission clusters (here named *TCs*). In case of nested clusters, mapping preferred the cluster corresponding to the most external node. Men included in the analyses also were required to have at least two years of follow-up with respect to semi-annual questionnaires of sexual behaviour. **Figure 2** illustrates this process with hypothetical versions of such pair of clusters. It depicts 2 behavioural clusters (black and light blue in **Figure 2A**) and 3 transmission clusters contained in a virus phylogeny (**Figure 2B**). The subsequent mapping is illustrated by **Figure 2C**. In this hypothetical example, Figure 2C suggests assortativity between members of the light blue behavioural cluster along transmission cluster *TC1*. This manuscript is concerned with the quantification of these patterns along the full virus phylogeny and the search for hints on potential associations between the two types of clustering (transmission versus sexual behaviour).

**Figure 2.**
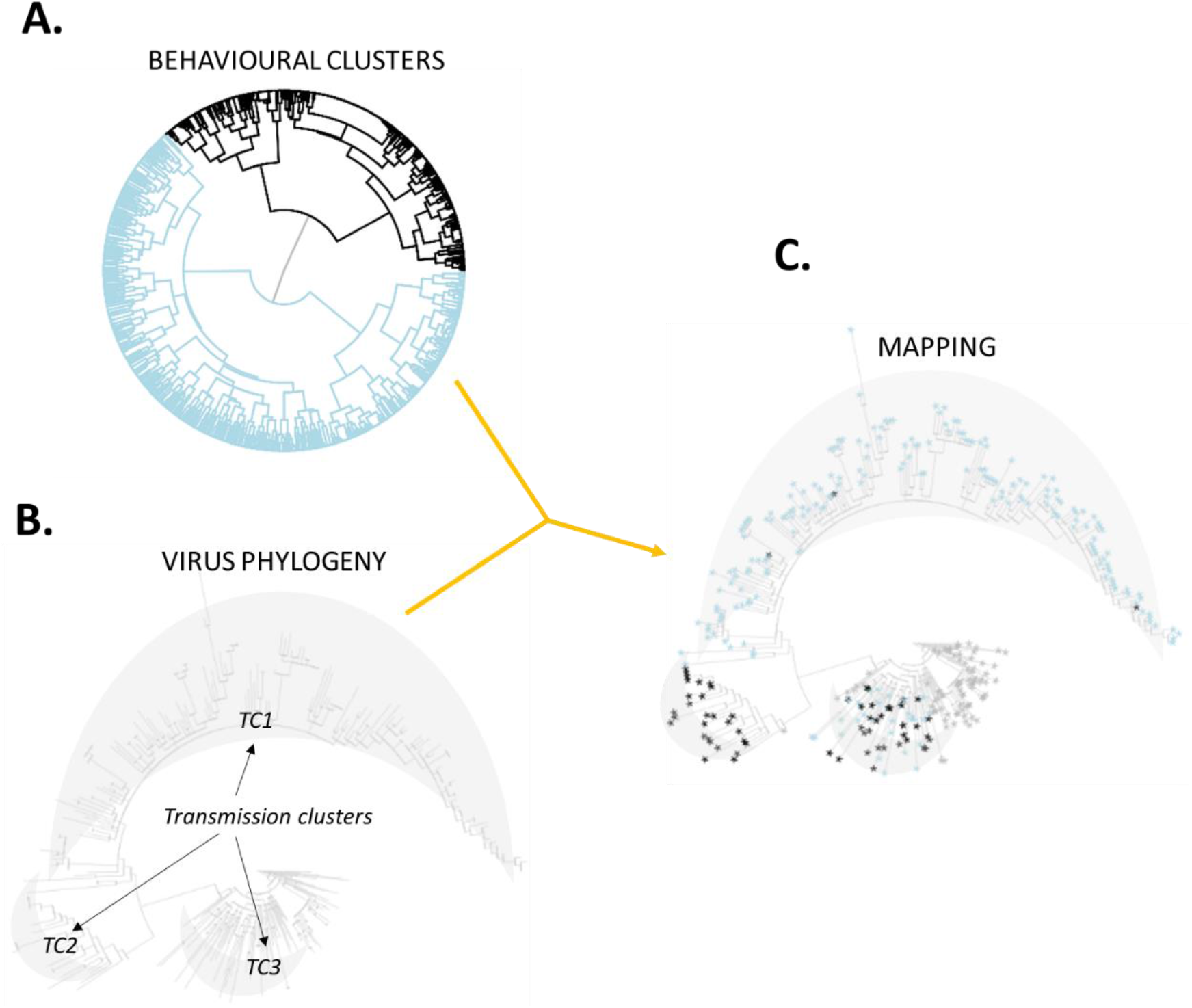
Hypothetical illustration of the mapping process. This example involves two behavioural clusters (panel A; light blue and black) and three transmission clusters (TC1, TC2 and TC3 in **panel B**). The mapping shown in **panel C** suggests assortativity among members of the light blue behavioural cluster in transmission cluster TC1 (containing only 2 members of the black behavioural cluster). Analogously, transmission cluster TC2 is exclusively formed by members of the black behavioural cluster (an unlikely observation if mixing was at random). Members of both behavioural clusters distribute evenly along transmission cluster TC3.

### Are MSM who share a behavioural cluster likely to also share transmission cluster?

To assess whether this was the case, we implemented a toy Monte Carlo analysis that iteratively shuffles transmission cluster membership at random. In each iteration it compared the fraction of independent pairs of men who share transmission cluster given that they share behavioural clusters with the fraction of pairs of men who do not share transmission clusters given that they share behavioural clusters. Outcomes were odds ratios and their corresponding p-values.

### Strength of interaction between behavioural clusters (Toy Monte Carlo)

We intended to establish a measure of how prone are members of behavioural clusters are to interact with each other over the whole set of HCV transmission clusters. For that purpose, let us assume that the distribution of behavioural cluster membership of sampled Swiss HCV sequences in each transmission cluster was generalizable, and mixing within HCV transmission clusters was homogeneous. Let us also assume interactions involving practices that could lead to HCV transmission occurring take place between MSM living in Switzerland. Given these assumptions, the probability over the whole HCV phylogeny that a member of behavioural cluster *BC*_*i*_ interacts with a member of behavioural cluster *BC*_*j*_ would be given by 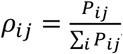, where 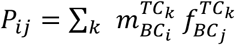, is our measure of interaction between these clusters, 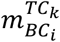 is the number of members of *BC*_*i*_ in *TC*_*k*_ and 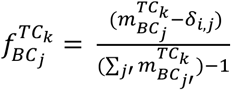 aims to reflect how common each behavioural cluster is in each transmission cluster. We used *ρ*_*ij*_ to quantify the strength of mixing between behavioural clusters *BC*_*i*_ and *BC*_*j*_.

### Potential influence of behavioural patterns on the HCV phylogeny (Interaction ratios)

We sought to identify over- or under-represented mixing, as well as assortativity between pairs of behavioural clusters. We did this by estimating the ratio *r*_*ij*_ between *ρ*_*ij*_ and its expected value if *TCs* membership was independent of *BC*s *i*.*e*., if behavioural clusters did not influence the conformation of HCV transmission clusters (or vice versa). If behavioural clusters did not influence HCV transmission clusters, the probability that a member of behavioural cluster *BC*_*i*_ interacts with a member of behavioural cluster *BC*_*j*_ would depend on the overall frequency of behavioural cluster *BC*_*j*_ along the HCV phylogeny, *i*.*e*., 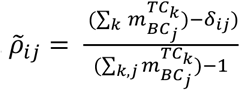. The ratio is therefore given by 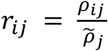. This ratio approaches 1 when HCV transmission clustering is independent of behavioural clustering, while ratios above 1 (potential over-representation) and below 1 (potential under-representation) could reflect an effect of behavioural groups on HCV transmission clustering. For this analysis, we defined over- and under-representation as ratios above two and below 0.5, respectively.

### Sample size and power considerations

In order to assess our ability to demonstrate a relationship between behavioural and transmission clusters, we estimated the number of individuals necessary to achieve a significance level of 0.05 and a power of 70% for the *toy Monte Carlo* and the *interaction ratios analyses*.

#### Toy Monte Carlo analysis

Experiments in this analysis were the number of independent pairs of men sharing TC. Independent pairs are those which cannot be inferred from other pairs. For example, if person *i* shares *TC* with persons *j* and *k*, then *j* and *k* also share TC. While it adds up to 3 pairs sharing cluster, only two would count as independent experiments.

#### The interaction ratios

These ratios were tested by assuming a stable structure of the intersection between the two types of cluster (i.e., the mapping). Therefore, that the *ratio* for a pair of *BC* required an increased number of members of these two clusters to reach a target level of significance, would also imply the need for increased numbers of members of all other clusters. This in order to guarantee that all *ratios* resemble the original ones.

These estimations relied on the R function *prop*.*test* and the R package *pwr[26]*. Because the hierarchical structure of the clustering allows for a shallower grouping (which naturally results in larger groups), we also applied all described methods to the two clusters resulting from the first bifurcation plus *BC*_*0*_. We termed these *three groups classification* and *full classification*. Panels **A** and **B** in **Figure 1** depict size and nsCAI trends in these groups.

## Data Availability

The individual level datasets generated or analyzed during the current study do not fulfill the requirements for open data access: 1) The SHCS informed consent states that sharing data outside the SHCS network is only permitted for specific studies on HIV infection and its complications, and to researchers who have signed an agreement detailing the use of the data and biological samples; and 2) the data is too dense and comprehensive to preserve patient privacy in persons living with HIV. According to the Swiss law, data cannot be shared if data subjects have not agreed or data is too sensitive to share. Investigators with a request for selected data should send a proposal to the respective SHCS address (www.shcs.ch/contact). The provision of data will be considered by the Scientific Board of the SHCS and the study team and is subject to Swiss legal and ethical regulations, and is outlined in a material and data transfer agreement.

## Author contribution statement

LSV and AR designed the study. LSV formulated and implemented the analyses, and drafted the first version of the manuscript, which was then revised by AR, KK and RDK. RDK contributed to data analyses and validation of results. HFG JB KM DLB DN EB AC, KEAD, GW and AR contributed to data collection and interpretation. HFG JB KM DLB DN MS EB AC, KEAD and GW contributed to interpreting the analyses and critically revised the manuscript.

## Acknowledgments

The authors thank all patients, physicians, and nurses associated with the SHCS, and to Andrew Atkinson for his suggestions.

## Disclaimer

The funders had no role in the study design, data collection and analysis, decision to publish, or preparation of the article.

## Financial support

This work was performed within the framework of the SHCS, supported by the Swiss National Science Foundation (SNSF grants 177499 and 324730_179567) and by the SHCS grant 823. D. L. B. was supported by the University of Zurich’s Clinical Research Priority Program Viral Infectious Diseases. R. D. K. was supported by the SNSF (grant BSSGI0_155851).

## Additional information

### Competing interests statement

L.S.V., K.K., D.N. and A.C. do not report competing interests.

H. F. G. has received unrestricted research grants from Gilead Sciences, Roche; a grant from Systems X; grants from SHCS research foundation, National Institutes of Health, Yvonne Jacob Foundation, personal fees from Data Safety Monitoring Board, and consulting for Merck, Gilead, Teva, and Sandoz, outside the submitted work.

A. R. reports support for advisory boards and/or travel grants from Janssen-Cilag, Merck Sharp & Dohme (MSD), Gilead Sciences, Abbvie, and Bristol-Myers Squibb, and an unrestricted research grant from Gilead Sciences. All remuneration went to his home institution and not to A. R. personally.

D. L. B. reports support for advisory boards and/or travel grants from MSD, ViiV, and Gilead Sciences.

R. D. K. reports personal fees from Gilead Sciences.

G. W. reports grants from Gilead Sciences and AbbVie.

K. E. A. D. reports contributions to her home institution from Gilead Sciences and grants from Merck Sharp and Dohme.

E. B. reports payments to his home institution from Gilead Sciences, MSD, ViiV Healthcare, Pfizer, and Sandoz, and grants from Gilead Sciences, MSD, ViiV, and Pfizer,

J.B. is a member of the Federal Commission for Issues relating to Sexually Transmitted Infections (CFIST);

K.J.M. has received travel grants and honoraria from Gilead Sciences, Roche Diagnostics, GlaxoSmithKline, Merck Sharp & Dohme, Bristol-Myers Squibb, ViiV and Abbott; and the University of Zurich received research grants from Gilead Science, Roche, and Merck Sharp & Dohme for studies that Dr Metzner serves as principal investigator, and advisory board honoraria from Gilead Sciences.

All these relationships were outside this work.

## SUPPLEMENTARY FIGURES

**Figure S1.**
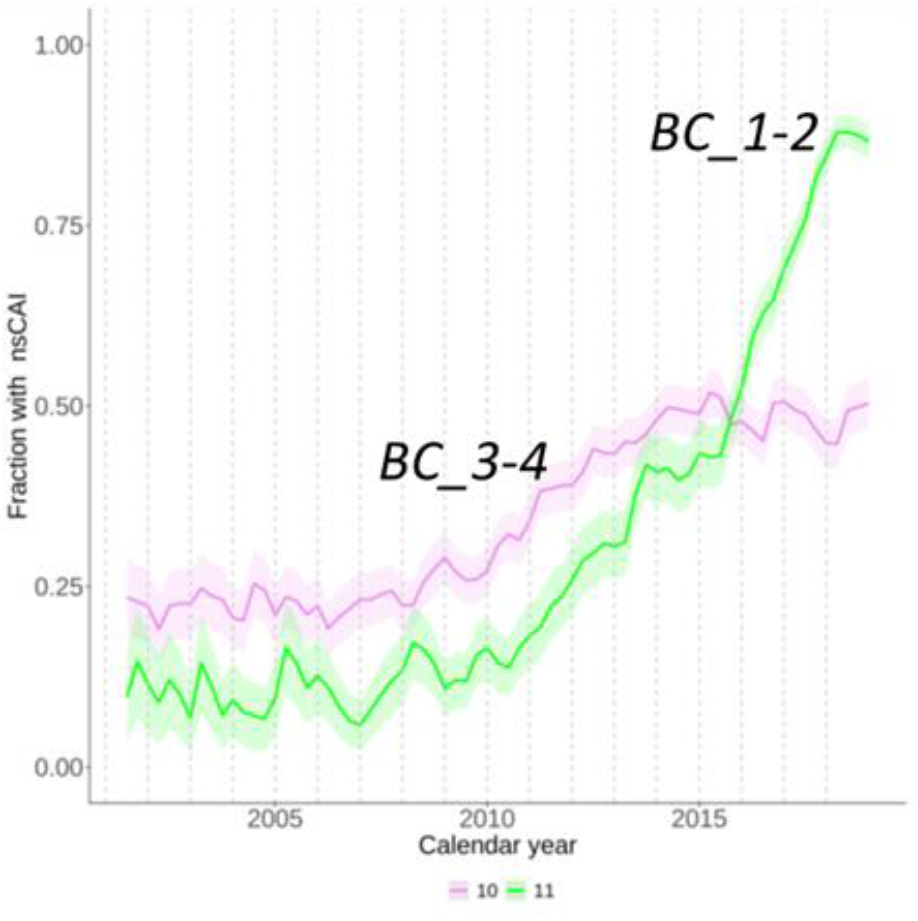
Trends in condomless anal intercourse with non-steady partners among MSM in the Three clusters classification. The analogous trends for the full classification are shown in [6].

## Notes

### Author Declarations

The SHCS has been approved by the ethics committee of the participating institutions (Kantonale Ethikkommission Bern, Ethikkommission des Kantons St. Gallen, Comite departemental d ethique des specialites medicales et de medicine communataire et de premier recours, Hopitaux Cantonale de Geneve, Kantonale Ethikkommission Zuerich, Repubblica e Cantone Ticino-Comitato Ethico Cantonale, Commission cantonale dêtique de la recherche sur l'être humain, Canton de Vaud, Lausanne, Ethikkommission beider Basel for the SHCS and Kantonale Ethikkommission Zuerich for the ZPHI). Written informed consent had been obtained from all participants. All methods were carried out in accordance with Swiss federal guidelines and regulations.

## REFERENCES

1. Hampel B, Kusejko K, Kouyos RD, et al. Chemsex drugs on the rise: a longitudinal analysis of the Swiss HIV Cohort Study from 2007 to 2017. HIV medicine 2020; 21(4): 228–39.

2. Kouyos RD, Hasse B, Calmy A, et al. Increases in Condomless Sex in the Swiss HIV Cohort Study. Open Forum Infect Dis 2015; 2(2): 4.

3. Nostlinger C, Platteau T, Bogner J, et al. Implementation and Operational Research: Computer-Assisted Intervention for Safer Sex in HIV-Positive Men Having Sex With Men: Findings of a European Randomized Multi-Center Trial. Journal of acquired immune deficiency syndromes 2016; 71(3): e63–72.

4. Sullivan PS, Carballo-Dieguez A, Coates T, et al. Successes and challenges of HIV prevention in men who have sex with men. Lancet 2012; 380(9839): 388–99.

5. Nostlinger C, Niderost S, Platteau T, et al. Sexual Protection Behavior in HIV-Positive Gay Men: Testing a Modified Information-Motivation-Behavioral Skills Model. Arch Sex Behav 2011; 40(4): 817–27.

6. Salazar-Vizcaya L, Kusejko K, Schmidt AJ, et al. Clusters of sexual behaviour in HIV-positive men who have sex with men reveal highly dissimilar time trends. Clinical infectious diseases : an official publication of the Infectious Diseases Society of America 2019.

7. Cohen MS, Chen YQ, McCauley M, et al. Antiretroviral Therapy for the Prevention of HIV-1 Transmission. The New England journal of medicine 2016; 375(9): 830–9.

8. Eisinger RW, Dieffenbach CW, Fauci AS. HIV Viral Load and Transmissibility of HIV Infection: Undetectable Equals Untransmittable. Jama 2019; 321(5): 451–2.

9. Rodger AJ, Cambiano V, Bruun T, et al. Risk of HIV transmission through condomless sex in serodifferent gay couples with the HIV-positive partner taking suppressive antiretroviral therapy (PARTNER): final results of a multicentre, prospective, observational study. Lancet 2019; 393(10189): 2428–38.

10. Vernazza, P., B Hirschel, E Bernasconi.,et al. “Les personnes séropositives ne souffrant d’aucune autre MST et suivant un traitement antirétroviral efficace ne transmettent pas le VIH par voie sexuelle.” Bulletin des médecins suisses| Schweizerische Ärztezeitung| Bollettino dei medici svizzeri 89.5 (2008): 165–169.

11. Salazar-Vizcaya L, Kouyos RD, Zahnd C, et al. Hepatitis C virus transmission among human immunodeficiency virus-infected men who have sex with men: Modeling the effect of behavioral and treatment interventions. Hepatology 2016; 64(6): 1856–69.

12. Andresen, S., S. Balakrishna, D. Nicca et al. “Behavioural patterns to identify key populations for syphilis prevention.” In HIV MEDICINE, vol. 20, pp. 219–219. 111 RIVER ST, HOBOKEN 07030-5774, NJ USA: WILEY, 2019.

13. Salazar-Vizcaya L, Kouyos RD, Metzner KJ, et al. Changing Trends in International Versus Domestic HCV Transmission in HIV-Positive Men Who Have Sex With Men: A Perspective for the Direct-Acting Antiviral Scale-Up Era. The Journal of infectious diseases 2019; 220(1): 91–9.

14. Rauch A, Rickenbach M, Weber R, et al. Unsafe sex and increased incidence of hepatitis C virus infection among HIV-infected men who have sex with men: the Swiss HIV Cohort Study. Clinical infectious diseases : an official publication of the Infectious Diseases Society of America 2005; 41(3): 395–402.

15. van Santen DK, van der Helm JJ, Del Amo J, et al. Lack of decline in hepatitis C virus incidence among HIV-positive men who have sex with men during 1990-2014. J Hepatol 2017; 67(2): 255–62.

16. Danta M, Rodger AJ. Transmission of HCV in HIV-positive populations. Current opinion in HIV and AIDS 2011; 6(6): 451–8.

17. Boerekamps A, van den Berk GE, Lauw FN, et al. Declining Hepatitis C Virus (HCV) Incidence in Dutch Human Immunodeficiency Virus-Positive Men Who Have Sex With Men After Unrestricted Access to HCV Therapy. Clinical infectious diseases : an official publication of the Infectious Diseases Society of America 2018; 66(9): 1360–5.

18. Braun DL, Hampel B, Kouyos R, et al. High Cure Rates With Grazoprevir-Elbasvir With or Without Ribavirin Guided by Genotypic Resistance Testing Among Human Immunodeficiency Virus/Hepatitis C Virus-coinfected Men Who Have Sex With Men. Clinical infectious diseases : an official publication of the Infectious Diseases Society of America 2018.

19. Bachmann N, Kusejko K, Nguyen H, et al. Phylogenetic Cluster Analysis Identifies Virological and Behavioral Drivers of HIV Transmission in MSM. Clinical infectious diseases : an official publication of the Infectious Diseases Society of America 2020.

20. Kusejko K, Marzel A, Hampel B, et al. Quantifying the drivers of HIV transmission and prevention in men who have sex with men: a population model-based analysis in Switzerland. HIV medicine 2018; 19(10): 688–97.

21. Amirkhanian YA. Social networks, sexual networks and HIV risk in men who have sex with men. Current HIV/AIDS reports 2014; 11(1): 81–92.

22. Basten M, Heijne JCM, Geskus R, Den Daas C, Kretzschmar M, Matser A. Sexual risk behaviour trajectories among MSM at risk for HIV in Amsterdam, the Netherlands. Aids 2018; 32(9): 1185–92.

23. Braun DL, Hampel B, Martin E, et al. High Number of Potential Transmitters Revealed in a Population-based Systematic Hepatitis C Virus RNA Screening Among Human Immunodeficiency Virus-infected Men Who Have Sex With Men. Clinical infectious diseases : an official publication of the Infectious Diseases Society of America 2019; 68(4): 561–8.

24. van Sighem A, Vidondo B, Glass TR, et al. Resurgence of HIV Infection among Men Who Have Sex with Men in Switzerland: Mathematical Modelling Study. PLoS One 2012; 7(9): 10.

25. Kohler P, Schmidt AJ, Cavassini M, et al. The HIV care cascade in Switzerland: reaching the UNAIDS/WHO targets for patients diagnosed with HIV. Aids 2015; 29(18): 2509–15.

26. Stephane Champely, Claus Ekstrom, Peter Dalgaard, Jeffrey Gill, Stephan Weibelzahl, Aditya Anandkumar, Clay Ford, Robert Volcic, Helios De Rosario; pwr: Basic Functions for Power Analysis. https://CRAN.R-project.org/package=pwr

